# Machine learning predicts relapse after anti-TNF withdrawal in ulcerative colitis using rectal biopsy transcriptomics

**DOI:** 10.64898/2026.10.05.26364793

**Authors:** Ghita Bouhddi, Emma Rapp, Piotr Artiemjew, Ingrid Prytz Berset, Frode Lerang, Dag Arne Lihaug Hoff, Espen Sønderaal Bækkevold, Knut E.A. Lundin, Frode Lars Jahnsen, Diana Domanska

## Abstract

**Background:** There are currently no available biomarkers that predict the clinical outcome of ulcerative colitis (UC) patients after anti-TNF withdrawal. Here we developed a semi-supervised machine learning framework on RNA-seq data derived from rectal biopsies to predict relapse following anti-TNF discontinuation.

**Methods:** We included UC patients (n=136) enrolled in the prospective, randomized controlled BIOSTOP trial. Patients were assigned to a withdrawal group (stopped treatment immediately) or a maintenance group (stopped treatment after 2 years). A machine learning framework was trained on gene expression data from mucosal biopsies collected at baseline from the withdrawal group and validated in the maintenance group using baseline samples (test cohort) and 2-year follow-up samples (interference cohort).

**Results:** A semi-supervised machine learning model, combining a shared UMAP manifold representation with LabelSpreading on a targeted 38-gene panel achieved an AUC of 0.794, a sensitivity of 90.3%, and a specificity of 73.1% on the test cohort, and an AUC of 0.711, a sensitivity of 82.9%, and a specificity of 42.3% on the inference cohort. Applying Shapley Additive exPlanations (SHAP) we found that *IL1B, TYROBP, REG1B, S100A8,* and *TNF* showed the highest predictive importance for relapse risk, whereas *MUC2, HMGCS2,* and *CDKN1A* displayed the highest predictive importance for sustained remission.

**Conclusion:** The high sensitivity makes the model a promising screening tool to prevent withdrawal-related relapse, though its moderate specificity underscores the need for multi-center validation in larger, more diverse cohorts before clinical deployment.

## Background and aims

Anti-tumor necrosis factor (anti-TNF) therapies are highly effective to induce long-lasting remission in patients with moderate-to-severe ulcerative colitis (UC)^1^. However, long-term treatment has side-effects, including increased infection risk, allergic reactions, and increased risk of some types of malignancies^2^. Moreover, long-term use may lead to loss of response due to production of anti-drug antibodies and has substantial healthcare costs^3^. The BIOSTOP trial is a prospective, multicenter, randomized, controlled, open-label study in which UC patients in clinical, biochemical, and endoscopic remission were randomized to either withdrawal or continuation of anti-TNF therapy. 53% of the patients in the withdrawal group relapsed within 2 years, which is consistent with other studies^4–8^. This suggests that anti-TNF can be safely de-escalated in a subset of patients who are in deep remission. However, at present, there are no reliable biomarkers that can identify which patients can safely stop treatment.

Although tissue transcriptomics offers a high-resolution window into residual mucosal inflammation, predicting disease trajectories from transcriptomic data presents a severe p >> n dimensionality problem: Thousands of genes are measured across a small cohort of patients. This imbalance creates a high risk of model overfitting, particularly given that transcriptional differences between subclinical remission and early relapse are subtle^9^. While machine learning has been broadly applied to inflammatory bowel disease (IBD) diagnosis and initial treatment response prediction^10–12^, models that forecast relapse in patients already meeting strict remission criteria remain scarce.

To overcome these limitations, we developed a semi-supervised machine learning framework tailored for small-sample transcriptomic datasets. The architecture combines unsupervised and supervised Uniform Manifold Approximation and Projection (UMAP) embeddings with graph-based label propagation using LabelSpreading. By pairing this framework with a literature-curated 38-gene panel, our approach achieves robust predictive generalization while retaining biological interpretability.

## Methods

### Patient cohort

Transcriptomic data were obtained from participants (n=163) enrolled in the BIOSTOP trial, a prospective, Norwegian multicenter, randomized controlled study evaluating the clinical outcome after anti-TNF withdrawal in patients with UC^13^. Trial eligibility required a confirmed UC diagnosis, continuous anti-TNF therapy for at least one-year, sustained clinical remission for at least 3 months, two consecutive fecal calprotectin measurements <200 µg/g, and endoscopic remission defined as Mayo endoscopic score (MES) 0 to 1. The study consisted of two parts. Part 1: 82 patients were randomized to discontinue anti-TNF therapy, while 81 continued maintenance treatment. Both groups were followed for 2 years with regular clinical assessments including mucosal sampling. Part 2: The patients were followed for additional 2 years. Here, 61 patients in the maintenance group stopped treatment. Detailed trial design and protocols have been published previously^13^.

The clinical study followed the principles of the Declaration of Helsinki and was approved by the Regional Committee for Medical Research Ethics (REK Midt 2016/531). All biopsies were deposited in the general biobank “Tarmsykdommer” (REK Sør-Øst #20521). Written informed consent was obtained from participants for both participation in the clinical study and storage of biological material in the general biobank.

Clinical parameters evaluated as potential covariates included age, sex, 6-point Mayo score (a composite disease severity index ranging from 0 to 6 combining stool frequency and rectal bleeding)^14^ MES (measuring endoscopic mucosal inflammation from 0 [inactive] to 3 [severe])^15^ and the Robarts Histopathology Index (RHI; a validated histological score quantifying leukocyte infiltration and epithelial damage)^16^.

### RNA sequencing

In this study we used bulk RNA-seq data of rectal biopsies collected at inclusion of the study and after 2 years follow-up. The samples were immediately immersed in RNAprotect Cell Reagent (Qiagen) and fresh frozen to -80°C prior to processing. RNA extraction, library construction, sequencing, and primary processing followed methods described previously^17^. Total RNA was extracted from frozen rectal biopsies using TRIzol reagent® from (InvitrogenTM: Cat. No.: 15596026) and the RNeasy Mini Kit (Qiagen: Cat. No.: 74106). Sequencing libraries were prepared using a stranded mRNA protocol and sequenced on an Illumina NovaSeq 6000 platform using paired-end 150 bp reads. Approximately 30 million read pairs were generated per sample (Novogene, UK).

Raw sequencing files were processed through the nf-core/rnaseq pipeline^18^ (v3.12.0), incorporating comprehensive quality control, transcript quantification and alignment. Key processing steps include adapter trimming via Trim Galore^19^, alignment to GRCh38 using STAR^20^, and quantification with Salmon^21^. Samples displaying high duplication rates (>70%), indicating low library complexity or excessive PCR amplification, were excluded.

### Training and test cohorts

Training cohort consisted of RNA-seq data from baseline biopsies of the anti-TNF withdrawal arm (n=72). We used samples from the maintenance group at baseline (n=57) as the independent test cohort and those at 2 years (n=61) as the inference cohort. Given that all maintenance participants received anti-TNF therapy prior to baseline randomization, both the baseline and 2-year follow-up samples provided biologically comparable controls for the baseline withdrawal group—differing primarily by two additional years of continued anti-TNF exposure.

### Preprocessing of RNA-seq data for machine learning

Raw count matrices were processed using the same four-step procedure in all cohorts. To ensure a strictly unbiased evaluation, all preprocessing parameters were estimated exclusively from the training cohort and subsequently applied unchanged to the independent test and interference cohorts.

The first step consisted of feature intersection. Only genes that were present in both the training and test datasets were retained. Next, variance stabilization step, a log1p transformation was applied to raw counts. This transformation reduces the effect of very large expression values, stabilizes the variance between genes, and prevents highly expressed genes from dominating the cosine-distance calculations used in UMAP. Next, gender correction step, where gender was evaluated as a potential biological confounder. For each gene, a separate linear regression model was fitted using gender as the explanatory variable in the training cohort. The estimated gender-related component was then removed from both the training and test datasets. The regression coefficients were estimated only from the training cohort and were applied unchanged to the test cohort. Finally, standardization step, the genes were standardized using a StandardScaler fitted only to the training cohort. Each gene was centered around a mean of zero and scaled to unit variance. The same scaling parameters were then applied to the test cohort.

## Results

### Patient demographics and clinical covariate selection

Clinical characteristics of patients included in this study are given in Table 1. Although the training and test cohorts were derived from the same clinical study, we evaluated whether global gene expression profiles exhibited cohort-specific shifts or technical treatment-arm biases. PCA revealed marked spatial overlap across both treatment strategies (maintenance vs. withdrawal) and clinical outcomes (Figure 1). PERMANOVA on the sequencing data confirmed no significant shift in global transcriptomic space between cohorts (pseudo-F = 0.89, p = 0.64), demonstrating that baseline expression profiles were statistically indistinguishable. Because clinical outcome variance was partially intertwined with cohort structure applying standard unconstrained batch-correction algorithms such as ComBat or removeBatchEffect from limma risked stripping away true biological signal alongside subtle technical noise. For this reason, we used a semi-supervised approach that could account for the difference between the cohorts without assuming a purely technical batch artifact.

**Figure 1.**
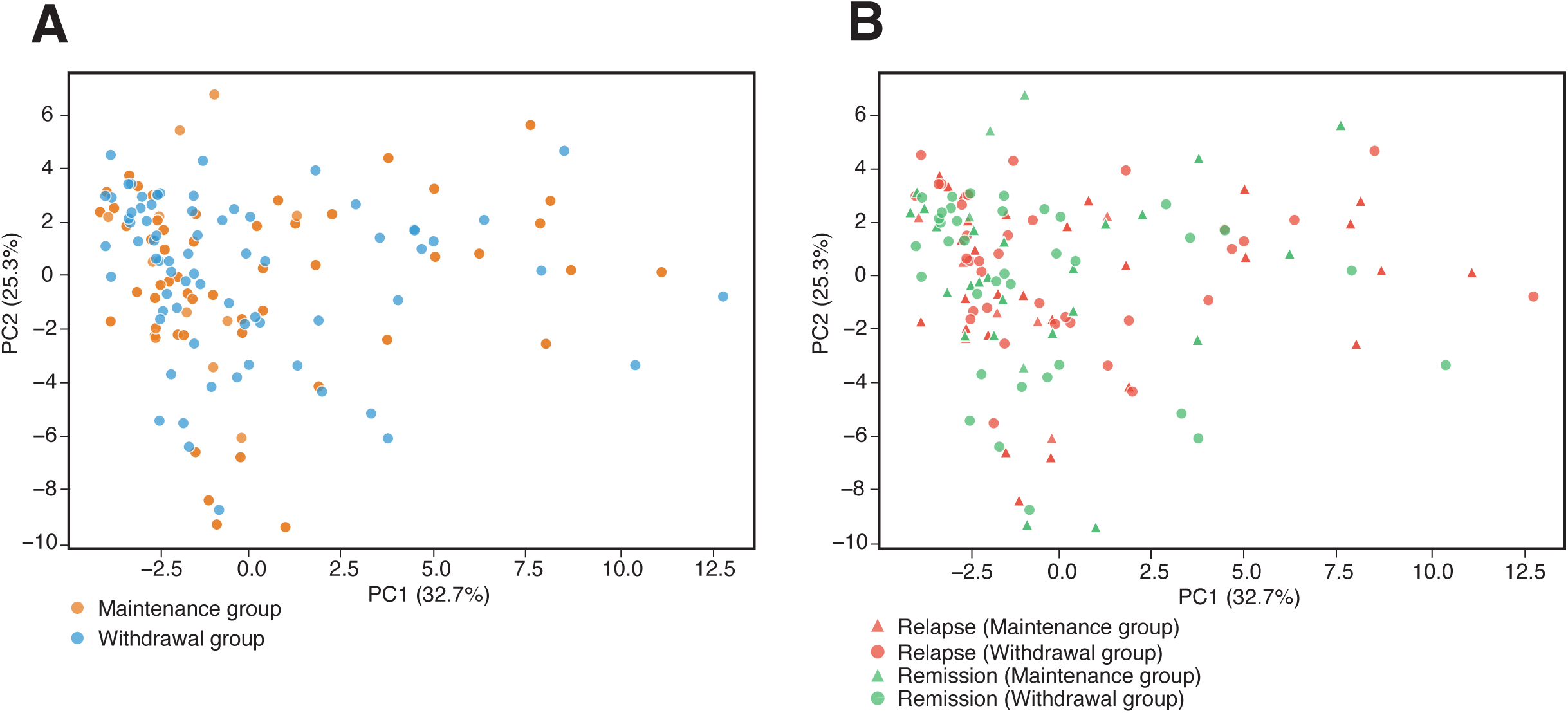
Principal Component Analysis of global transcriptomic profiles of training and testing cohorts. **(A)** PCA score plot colored by treatment group (Maintenance group in orange; Withdrawal group in blue), illustrating global transcriptomic homogeneity across treatment arms. **(B)** PCA score plot colored by clinical outcome and treatment strategy interaction, showing overlapping distributions among Relapse (red) and Remission (green) across both Maintenance (triangles) and Withdrawal (circles) cohorts.

**Table 1.**
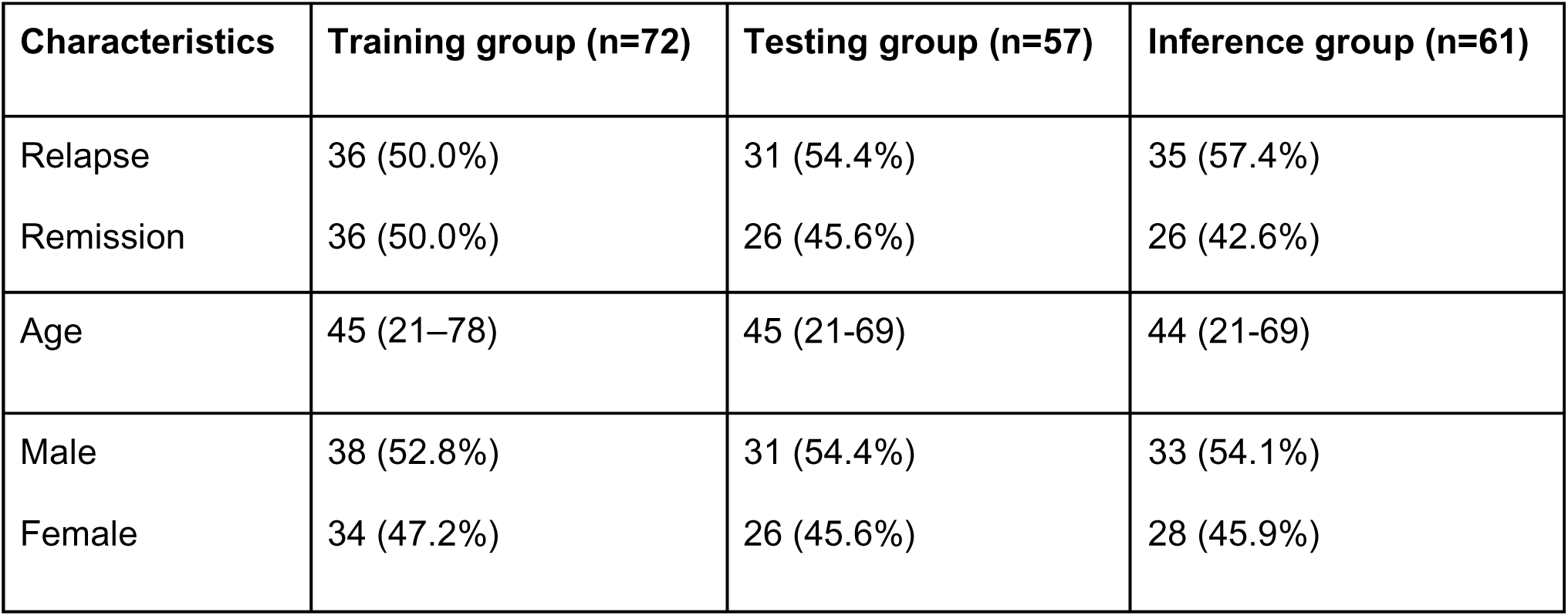
Baseline demographic and clinical characteristics of patients in the relapse and remission group.

We further evaluated whether incorporating clinical covariates into the preprocessing pipeline could enhance predictive performance. Six candidate variables were tested: gender, age, baseline Mayo score, RHI, RIN, and sequencing batch. Each variable was benchmarked as a preprocessing covariate using the semi-supervised classification pipeline. Gender was the only variable that consistently improved model performance; omitting gender-based correction reduced balanced accuracy from 0.805 to 0.773. This confirmed that adjusting for gender-specific expression variation minimized confounding and improved generalization to the independent test cohort. In contrast, inclusion of age, Mayo score, RHI, RIN, or batch effects did not improve classification performance, and degraded model performance across several configurations. Gender was therefore retained as the sole clinical covariate in the final preprocessing workflow.

### Feature selection strategy and optimization of the gene panel

The preprocessed training dataset contained 17,807 genes across 72 samples. We initially implemented a data-driven feature-selection strategy, evaluating differential expression and feature importance across 50 stratified bootstrap resamples using seven complementary metrics: FDR-corrected differential expression, log2 fold-change directionality, Cohen’s d effect size, Random Forest permutation importance, logistic regression coefficients, SHAP, and LIME. However, constrained sample size introduced substantial feature instability, yielding 1,040 unique genes among the top-ranked candidates across bootstrap iterations. This high variance demonstrated that purely data-driven selection was insufficient to establish a reproducible feature set.

To mitigate overfitting and construct a robust predictive feature space, we systematically evaluated varying panel sizes and modeling architectures. Initial feature sets of 100, 90, 70, 50, and 40 genes were benchmarked using a composite-scoring framework paired with a Label Spreading semi-supervised pipeline. Predictive performance steadily improved as the panel was refined to a smaller, biologically targeted core, with the area under the curve (AUC) increasing from 0.698 to 0.721 (Table 2). Incorporating non-linear manifold learning (UMAP) initialized on a literature-curated 20-gene anchor set^22–26^ yielded a marked step-change in model performance (AUC = 0.753). The feature space was subsequently expanded to 45 genes by blending literature-validated targets with computationally ranked candidates, followed by final optimization to a 38-gene panel that combined literature curation with unsupervised and supervised UMAP dimensionality reduction. This 38-gene configuration demonstrated the strongest predictive capacity across all development stages (F1 remission increased from 0.727 to 0.792; F1 relapse increased from 0.720 to 0.848; AUC increased from 0.748 to 0.794).

**Table 2.** Iterative refinement of the transcriptomic panel and machine learning architecture across model revisions.

| Revision | Steps | Genes panel size | F1 remission | F1 relapse | AUC | Balanced accuracy |
| --- | --- | --- | --- | --- | --- | --- |
| 1 | Initial 4-dimensional composite score with Label Spreading | 100 | 0.694 | 0.681 | 0.698 | 0.695 |
| 2 | Label Spreading gamma tuning | 90 | 0.701 | 0.692 | 0.702 | 0.701 |
| 3 | Included gender as covariate | 90 | 0.712 | 0.703 | 0.710 | 0.708 |
| 4 | StandardScaler with log1p | 70 | 0.718 | 0.707 | 0.715 | 0.712 |
| 5 | Replaced composite score with PCA | 70 | 0.704 | 0.698 | 0.709 | 0.704 |
| 6 | Switched to cosine distance | 50 | 0.715 | 0.710 | 0.718 | 0.714 |
| 7 | Balanced-accuracy threshold instead of F1 threshold | 40 | 0.724 | 0.718 | 0.718 | 0.721 |
| 8 | Alpha parameter tuning (0.7) | 40 | 0.728 | 0.720 | 0.721 | 0.724 |
| 9 | 5-fold cross-validated threshold optimization | 40 | 0.731 | 0.725 | 0.721 | 0.728 |
| 10 | UMAP using literature-selected genes | 20 | 0.738 | 0.734 | 0.753 | 0.740 |
| 11 | Early UMAP parameter tuning | 20 | 0.678 | 0.671 | 0.712 | 0.675 |
| 12 | Unsupervised UMAP on combined samples | 45 | 0.727 | 0.720 | 0.748 | 0.733 |
| 13 | Blended unsupervised (0.9) and supervised (0.1) UMAP | 38 | 0.793 | 0.786 | 0.771 | 0.797 |
| 14 | 0.92 unsupervised and 0.08 supervised UMAP with MCC threshold | 38 | 0.792 | 0.848 | 0.794 | 0.817 |

To ensure biological grounding, candidate genes in the final panel were cross-referenced against expression signatures from published UC studies^22–26^. The final panel contained 38 genes: AQP8, CA1, CA2, CDKN1A, CEACAM7, CLCA1, CYP3A4, CXCL1, DUOX2, DUOXA2, FABP1, FABP2, FCGR3B, GUCA2A, GUCA2B, HMGCS2, IL1B, IL6, LCN2, MS4A12, MMP3, MMP10, MUC2, MUC4, OSM, OSMR, REG1A, REG1B, REG3A, S100A8, S100A9, SLC26A2, SLC26A3, SLC39A4, SPI1, TNF, TREM1, TYROBP.

### Machine learning pipeline architecture

The classification framework integrated manifold learning with graph-based label propagation using the optimized 38-gene panel. High-dimensional expression profiles were projected into a hybrid coordinate space formed by linearly combining an unsupervised UMAP embedding of all samples (weight=0.92) with a supervised UMAP embedding fitted exclusively on the labeled training cohort (weight=0.08).

Hyperparameters for the unsupervised UMAP embedding were set to n_components = 5, min_dist = 0.1, n_neighbors = 15, and cosine distance. For the supervised UMAP embedding, parameters were tuned to n_components = 3, min_dist = 0.3, n_neighbors = 10, metric = cosine. The blended embedding matrix was subsequently classified using Label Spreading with a Radial Basis Function (RBF) kernel (γ = 1.0, α = 0.7). In this semi-supervised graph setup, training samples served as labeled nodes to propagate continuous relapse probabilities to unlabeled test nodes.

To determine classification decision thresholds without introducing data leakage, 5-fold out-of-fold (OOF) cross-validation was conducted strictly within the training set to maximize the Matthews Correlation Coefficient (MCC). This yielded an OOF-optimized threshold of 0.450 for the transductive evaluation path, and 0.430 for the inductive path used to predict unseen samples. No outcomes from the independent test or 2-year inference cohorts were used for threshold selection. Model calibration was assessed via the Hosmer-Lemeshow goodness-of-fit test. Primary evaluation metrics included sensitivity, specificity, overall accuracy, precision, recall, class-specific F1-scores, balanced accuracy, and area under the receiver operating characteristic curve (AUC). The independent test cohort was held out and isolated during feature selection, hyperparameter optimization, and threshold tuning.

### Performance in the testing cohort

Evaluated on the independent test cohort, the semi-supervised model demonstrated strong predictive power and cross-cohort generalization (AUC = 0.794, balanced accuracy= 0.817; Figure 2, Supp figure 1). Outcome-specific discrimination was well-balanced, yielding F1-scores of 0.848 for relapse and 0.792 for remission. At the OOF-optimized decision threshold, the model prioritized high sensitivity (0.903) alongside moderate specificity (0.731), successfully identifying 90.3% of patients who relapsed following anti-TNF withdrawal (Figure 2A). This sensitivity-focused calibration aligns with the primary clinical objective of minimizing false-negative predictions to identify patients at high risk of disease flare before therapy discontinuation.

**Figure 2.**
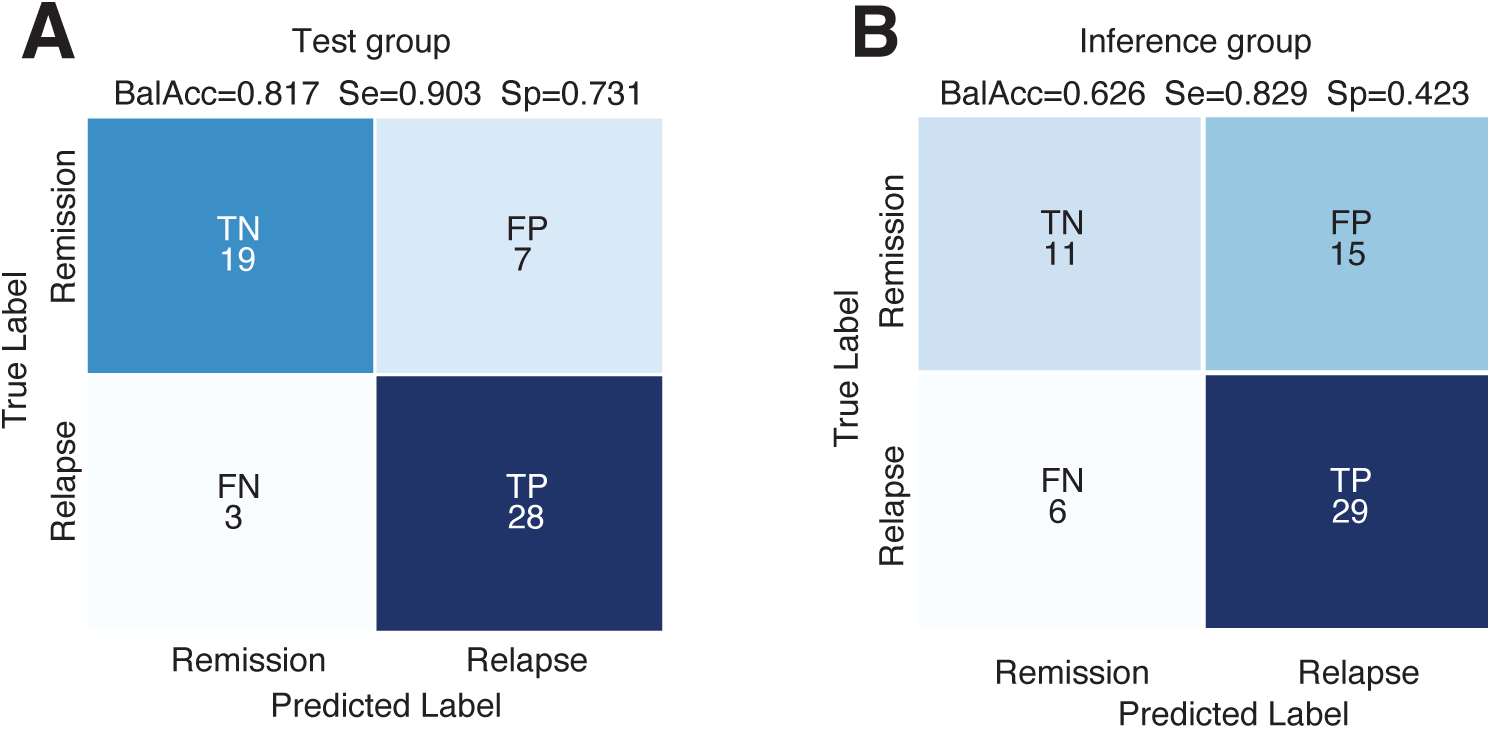
Classification performance of the 38-gene transcriptomic model in test and inference cohorts. **(A)** Confusion matrix showing classification counts for relapse and remission in test group A at the out-of-fold (OOF)-optimized decision threshold maximizing the Matthews correlation coefficient (MCC). **(B)** Confusion matrix showing classification counts for relapse and remission in inference group at the same decision threshold.

### Longitudinal inference on maintenance group

To evaluate model performance over time, the baseline-trained classifier was evaluated on the independent 2-year continuous maintenance cohort. The model retained substantial predictive capability (AUC = 0.711, F1 relapse = 0.734, decision threshold = 0.430). High sensitivity was largely preserved (0.829), confirming that core inflammatory risk signatures remain detectable despite extended treatment. However, model specificity decreased to 0.423 (balanced accuracy= 0.626, F1 remission= 0.512) (Figure 2B).

To evaluate whether the drop in longitudinal specificity stemmed from a global distributional shift in model predictions, we examined changes in predicted relapse probabilities between baseline and 2-year follow-up. In paired Wilcoxon signed-rank analyses of repeated patient measurements, patients who experienced a relapse showed a slight increase in predicted relapse probability from baseline to the 2-year follow-up (n=30, median change=0.007, p=0.0345). Conversely, baseline samples from patients who remained in sustained remission (n=24) showed no significant change in predicted probability (median change≈0, p=0.7469). Descriptive distribution analyses further supported overall score stability (relapse: Kolmogorov–Smirnov [KS] p=0.891, Population Stability Index [PSI]=0.096; remission: KS p=0.733, PSI=0.162).

We next evaluated whether choice of decision threshold contributed to this effect. Applying the transductive threshold of 0.450 (rather than the inductive threshold) to 2-year predictions increased specificity from 0.423 to 0.577 (false positives dropping from 15 to 11) while marginally reducing sensitivity from 0.829 to 0.771 (false negatives increasing from 6 to 8).

Together, these findings indicate that while the lower inductive threshold partially contributed to lower specificity at 2 years, the primary driver is a subtle class-overlap shift near the decision boundary under extended anti-TNF exposure, rather than a broad collapse of score distributions (Figure 3, Supp figure 2).

**Figure 3.**
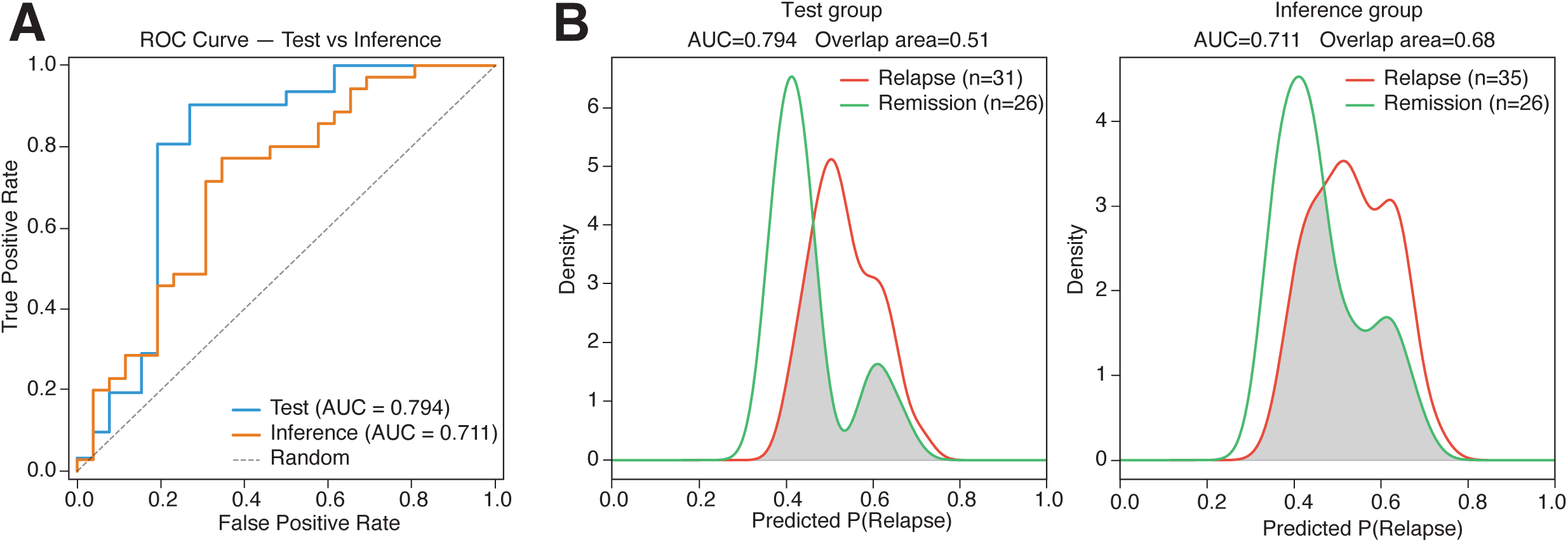
Performance of model across test and inference groups. **(A)** ROC curves comparing the model’s performance on the baseline test cohort (blue line) versus the 2-year longitudinal maintenance inference cohort (orange line). **(B)** Class separation density plots for test evaluation. **(C)** class separation density plot for inference evaluation.

### Permutation feature importance and biological interpretation

Permutation-based feature importance analysis (AUC = 0.777, n repeats = 5) identified key predictive drivers in the 38-gene panel (Figure 4A). Permutation of inflammation-related markers (e.g., *TREM1, CXCL1, OSM, MMP10*) induced the largest drops in classification performance, alongside essential epithelial barrier markers (e.g., *CDKN1A, HMGCS2)*.

**Figure 4.**
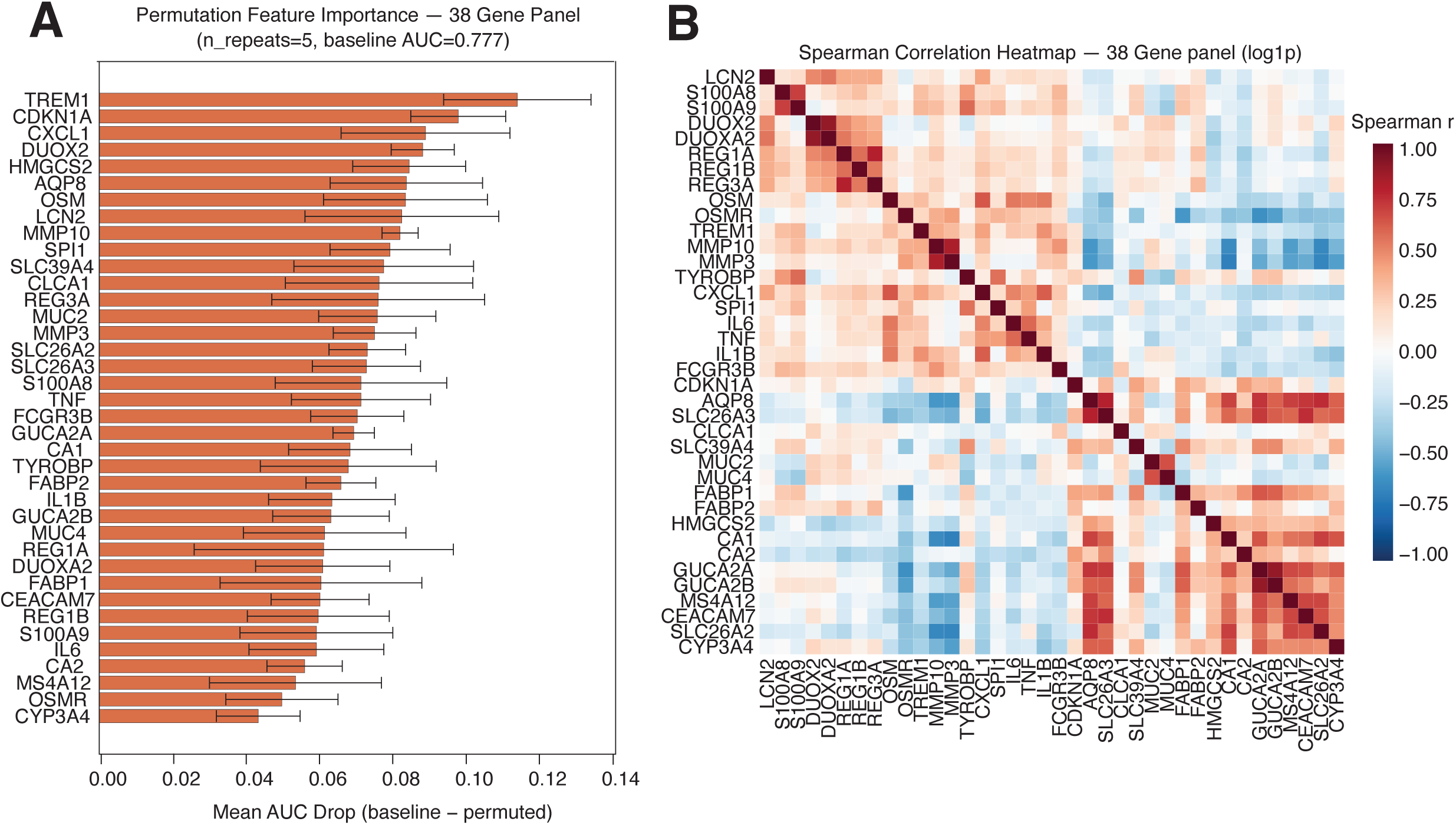
Feature importance and co-expression landscape of the 38-gene transcriptomic panel. **(A)** Permutation feature importance ranking the top predictive features by mean AUC drop. Error bars represent standard deviation across permutation repeats. **(B)** Spearman correlation heatmap displaying pairwise expression relationships across all 38 panel genes in n = 129 mucosal biopsy samples (log1p transformed). Color intensity reflects the Spearman correlation coefficient.

Spearman correlation analysis across the 38 features revealed two distinct co-expression modules, showing strong positive correlations among genes associated with immune activity and inflammation, and separate, tight clustering among genes that are associated with epithelial barrier functions (Figure 4B). Notably, strong reciprocal anti-correlation (blue regions) was observed between the modules, illustrating a clear transcriptional trade-off between inflammation-related activity and preserved epithelial barrier integrity.

### SHAP analysis dissects individual gene contributions and directionality

To translate model mechanics into clinically actionable biological insights, we applied SHapley Additive exPlanations (SHAP) across the independent testing cohort.

Global importance ranking established the proinflammatory cytokine *IL1B* as the single most influential driver of model decisions (mean |SHAP|=0.011). This was followed by genes associated with functions such as tissue remodeling (*MMP3*), antimicrobial activity (*REG1B, S100A8*), and metabolic activity *CYP3A4* (mean |SHAP| around 0.003-0.005, Figure 5A).

**Figure 5.**
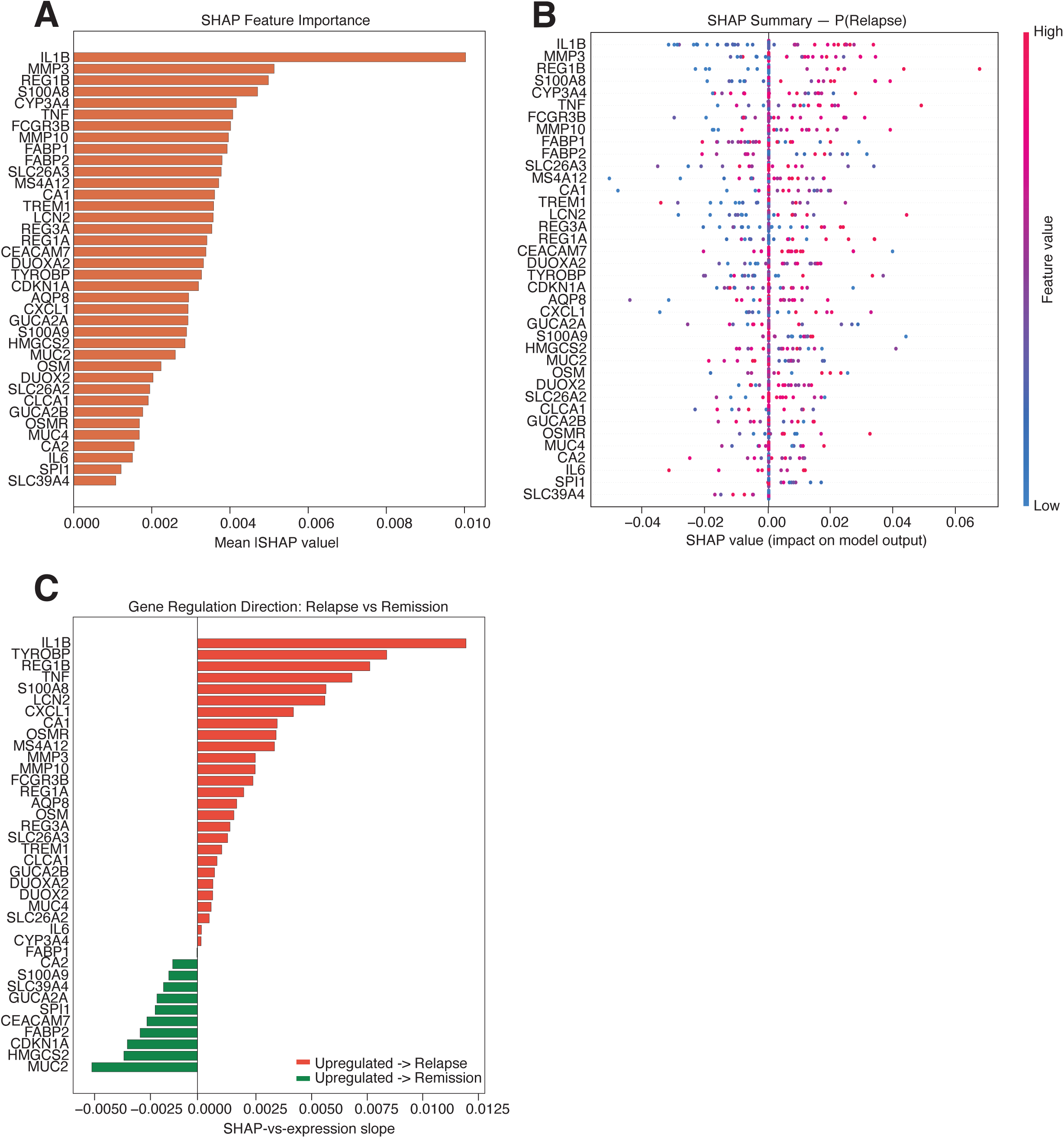
SHAP explainability analysis translating gene expression to clinical relapse risk (n = 57). **(A)** Global feature ranking displaying top predictive genes ordered by overall model impact (mean |SHAP| values. **(B)** SHAP beeswarm plot demonstrating how individual gene expression levels shift predicted relapse risk. Each dot represents an individual patient biopsy; color denotes relative transcript abundance (red: high; blue: low). Points extending right of zero indicate an increased probability of relapse following anti-TNF withdrawal. **(C)** Gene regulation direction of top predictive genes, red bars indicate that it is upregulated in relapse, and green indicates that it is upregulated in remission

Analysis of individual sample predictions via the SHAP summary plot revealed a clear biological trade-off between inflammation-related activity and tissue homeostasis. Specifically, high feature value of inflammation-related markers (e.g., *IL1B, MMP3, REG1B, S100A8*) systematically pushed the model outputs toward an increased predicted risk of relapse. In contrast, high expression of genes governing epithelial barrier integrity (e.g., *FABP1, MUC2, SLC39A4*) exerted a protective effect, shifting predictions toward sustained remission (Figure 5B).

Finally, to identify key drivers of clinical outcomes, SHAP-vs-expression slope analysis was utilized to evaluate the directional impact of gene regulation on relapse risk. Upregulation of markers such as *IL1B*, *TYROBP*, *REG1B, and TNF* demonstrated positive slopes driving predicted relapse risk upward (27 in total). Conversely, genes including *MUC2*, *HMGCS2*, and *CDKN1A* displayed negative SHAP slopes, indicating that their elevated expression is associated with clinical remission (11 in total) (Figure 5C).

## Discussion

Here we developed and independently validated a semi-supervised machine learning framework that pairs UMAP and LabelSpreading with a 38-gene panel to forecast relapse following anti-TNF discontinuation of UC patients in deep remission. Evaluated on an independent test cohort, the model demonstrated strong predictive power (AUC = 0.794, balanced accuracy= 0.817), capturing relapse risk with high sensitivity (0.903) and robust outcome-specific precision (F1 relapse = 0.848, F1 remission = 0.792). Furthermore, evaluating the model across a longitudinal continuous maintenance cohort confirmed sustained discrimination (AUC = 0.711, balanced accuracy= 0.626, sensitivity= 0.829), illustrating that core molecular risk signatures remain detectable despite ongoing anti-TNF exposure.

A central finding of this study is that a compact, biologically targeted transcriptomic panel provides robust predictive capacity in a clinical setting where conventional gene-by-gene differential expression analysis failed to identify individual markers reaching genome-wide statistical significance^17^. This divergence underscores that post-withdrawal disease trajectory is governed by subtle, higher-order gene co-expression networks rather than single-gene expression spikes. By shifting from univariate hypothesis testing to multivariate graph-based learning, our framework successfully extracts distributed, network-level signals associated with subclinical disease activity.

We used SHAP analysis to give biological insights into our machine learning model. Importantly, we found that the genes contributing to relapse prediction were all known to be involved in the inflammatory process of UC such as *S100A8, LCN2, OSM, TREM1, IL1B, IL6, TNF,* and *CXCL1* ^27,28^, whereas genes contributing to remission were predominantly related to the maintenance of a well-functioning epithelial barrier (e.g., MUC2, HMGCS2, FABP2, CDKN1A, and CEACAM7)^29–33^.

In small-sample transcriptomic studies (p >> n), standard data-driven feature selection is prone to severe overfitting. Our initial bootstrap-based feature selection yielded over 1,000 unstable candidate genes across resamples. Incorporating prior biological knowledge from literature^22–26^ stabilized the feature space, reducing dimensionality while preserving biological interpretability. Additionally, gender emerged as the sole clinical covariate that consistently improved model performance (increasing balanced accuracy from 0.773 to 0.805). Correcting for gender-associated transcriptomic variation removed a major source of biological confounding without obscuring disease-relevant signal, whereas age, Mayo score, RHI, RIN, and sequencing batch did not enhance out-of-sample generalization.

Primary model testing was conducted on independent baseline biopsies prior to drug withdrawal, with prospective inference performed on the two-year continuous maintenance cohort. While the model maintained high sensitivity and stable prediction probabilities over time, specificity decreased at two years. Sensitivity analyses indicated this reduction was partially driven by operating threshold differences between the transductive and inductive paths: applying the 0.450 baseline threshold raised 2-year specificity from 0.423 to 0.577 (compared to 0.731 at baseline). Additionally, paired analyses showed stable relapse probabilities among remission patients, demonstrating that the specificity decline was not simply caused by systematic treatment-induced transcriptomic shifts.

In a treatment-withdrawal setting, avoiding false negatives is paramount to prevent disease reactivation upon drug discontinuation. The model demonstrated high sensitivity for relapse in both the baseline test cohort (0.903 at the transductive threshold of 0.450) and the 2-year inference cohort (0.829 at the inductive threshold of 0.430). However, this came at the cost of lower specificity, particularly at two years. Threshold sensitivity analysis illustrated this trade-off: applying the 0.450 transductive threshold to 2-year predictions increased specificity from 0.423 to 0.577, while sensitivity decreased from 0.829 to 0.771. Consequently, the model’s moderate specificity means a subset of stable patients may be advised to remain on anti-TNF therapy unnecessarily. Future work will focus on validating this 38-gene panel in external multi-center cohorts, refining clinical decision cutoffs, and integrating non-invasive biomarkers, such as fecal calprotectin, to enhance specificity without sacrificing sensitivity.

### Limitations

This study has several limitations. First, while substantial for prospective mucosal tissue transcriptomics, the overall cohort size (n=136 patients across training, testing, and longitudinal inference sets) remains modest, leaving the pipeline susceptible to residual sampling variation. Second, all participants were enrolled in the Norwegian multicenter BIOSTOP trial. Because this represents a geographically restricted population, external validation in larger, multi-center, and ethnically diverse cohorts is necessary to confirm generalizability. Third, transcriptomic profiling was restricted to rectal mucosal biopsies collected at single timepoints. Because ulcerative colitis is characterized by spatial and temporal heterogeneity, a single rectal sample may fail to reflect active disease in more proximal colonic regions or capture rapid molecular dynamics preceding a relapse. Finally, the balance between sensitivity and specificity in this pipeline reflects a specific operating threshold trade-off. Establishing calibrated decision cutoffs that minimize unnecessary treatment continuation while maintaining high safety margins will require validation in larger prospective cohorts.

## Conclusion

We developed and validated a biologically guided 38-gene transcriptomic framework combined with semi-supervised graph-based machine learning (UMAP and LabelSpreading) to predict relapse prior to anti-TNF discontinuation in UC patients in deep remission. The framework successfully captured coordinated network-level shifts where conventional univariate differential expression testing detected no significant single-gene markers, mapping a clear biological trade-off between subclinical mucosal inflammation and restored epithelial barrier integrity. Furthermore, our findings highlight the necessity of leveraging biological prior knowledge and strict cohort isolation to avoid overfitting in high-dimensional, small-sample transcriptomic studies. Although the model’s high sensitivity supports its potential as a clinical screening tool to prevent premature anti-TNF withdrawal, its moderate specificity and single-trial origin necessitate external validation before clinical deployment. Integrating this 38-gene signature with non-invasive biochemical, endoscopic, and histological markers offers a promising pathway toward safe, individualized anti-TNF withdrawal strategies.

**Supplementary figure 1.** ROC and Precision-Recall curves evaluating model performance on the **(A)** test group and **(B)** inference group.

**Supplementary figure 2. (A)** Histograms comparing predicted relapse probabilities for test (blue) versus inference (orange) datasets in true relapse (left) and remission (right) patients, including statistical distribution comparisons. **(B)** predicted relapse probability for each relapse patient (left) and remission patient (right) using Wilcoxon signed-rank test.

## Abbreviations used in the paper

AUC: Area under the curve
FDR: False discovery rate
IBD: Inflammatory bowel disease
KS: Kolmogorov–Smirnov
LIME: Local interpretable model-agnostic explanations
MCC: Matthews correlation coefficient
MES: Mayo endoscopic score
OOF: Out-of-fold
PCA: Principal component analysis
PERMANOVA: Permutational multivariate analysis of variance
PSI: Population Stability Index
RBF: Radial basis function
RHI: Robarts histopathology index
RIN: RNA integrity number
ROC: Receiver operating characteristic
SHAP: Shapley additive explanations
TNF: Tumor necrosis factor
UC: Ulcerative colitis
UMAP: Uniform manifold approximation and projection

## Data Availability

The sequencing data underlying the results of this analysis will be made available upon reasonable request to the corresponding author, subject to applicable ethical and data-protection requirements.

## Notes

### Competing Interest Statement

The authors have declared no competing interest.

### Author Declarations

Regional Committee for Medical Research Ethics (REK Midt 2016/531)

