## Supplementary figures and images for "Machine learning predicts relapse after anti-TNF withdrawal in ulcerative colitis using rectal biopsy transcriptomics"

### Supp figure 1

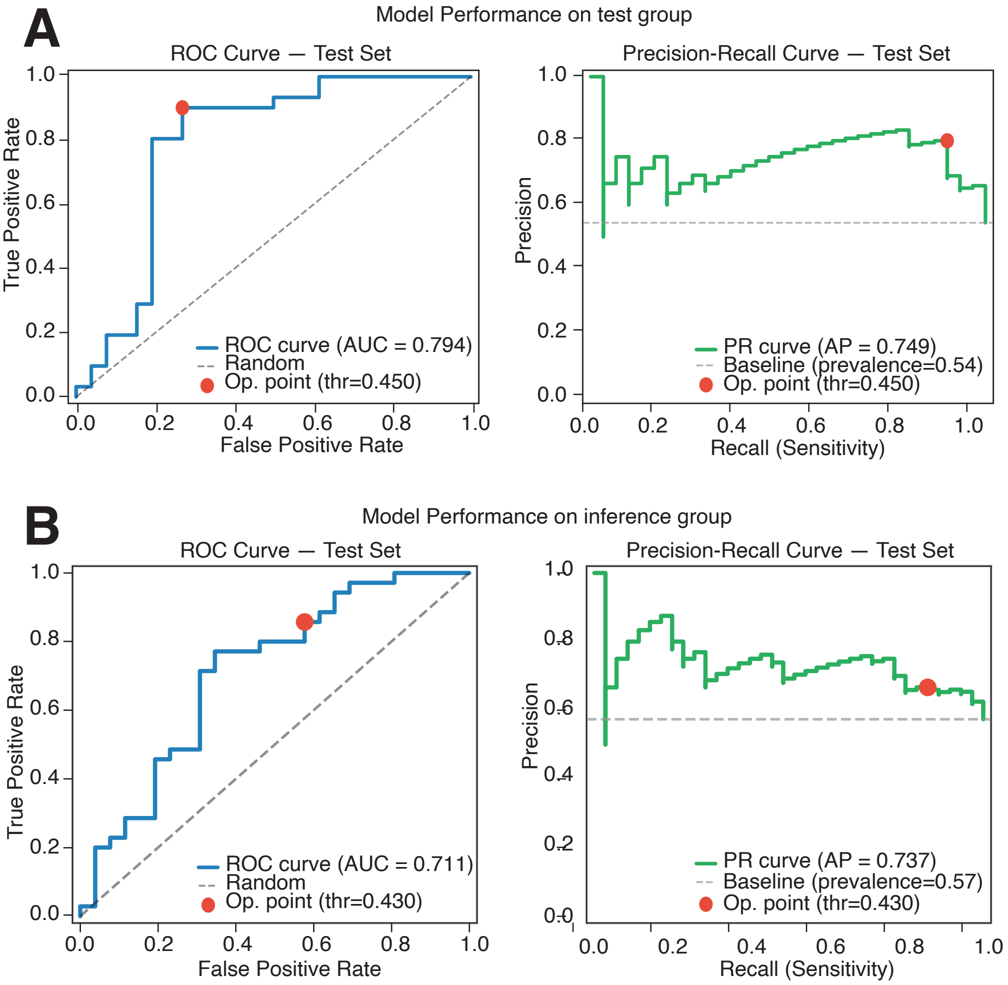

### Supp figure 2

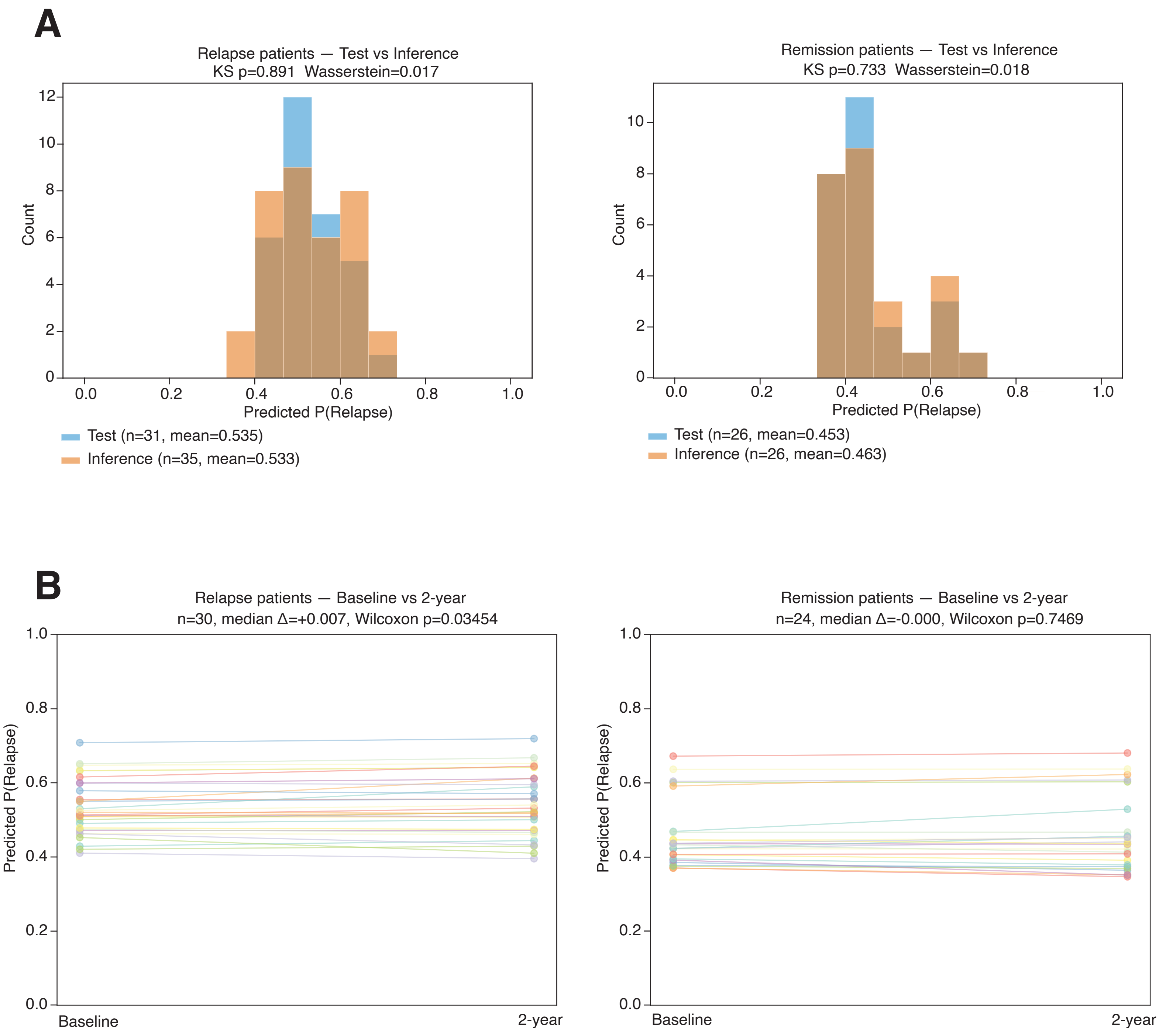
